# Highly heterogeneous human herpes virus-8 oral shedding kinetics among people with and without Kaposi sarcoma and HIV- co-infection

**DOI:** 10.1101/2024.05.17.24307135

**Authors:** Elizabeth M. Krantz, Innocent Mutyaba, Janet Nankoma, Fred Okuku, Corey Casper, Jackson Orem, David A. Swan, Warren Phipps, Joshua T. Schiffer

**Affiliations:** Vaccine and Infectious Disease Division, Fred Hutchinson Cancer Center, Seattle, USA; Uganda Cancer Institute, Kampala, Uganda; Access to Advanced Health Institute, Seattle, USA; Division of Allergy and Infectious Diseases, University of Washington, Seattle, USA

**Keywords:** HHV-8, viral shedding dynamics, Kaposi Sarcoma, HIV, KSHV

## Abstract

**Background:** An improved understanding of oral human herpesvirus-8 (HHV-8) viral dynamics could provide insights into transmission risk and guide vaccine development.

**Methods:** We evaluated HHV-8 oral shedding dynamics in Ugandan adults stratified by Kaposi sarcoma (KS) and HIV status. Participants were followed for ≥4 weeks, with daily home oral swab collection to quantify HHV-8 using Polymerase Chain Reaction. Shedding rates were defined by the number of days with HHV-8 detected divided by the total days with swabs and compared by group using hurdle models.

**Results:** 295 participants were enrolled; median age was 35 years (range 18-71), 134 (45%) were male. HHV-8 was detected more frequently among participants with KS (HIV+/KS+ 56/76, 74%; HIV-/KS+ 9/18, 50%) than those without KS (HIV+/KS-36/125, 29%; HIV-/KS-16/76, 21%); odds of shedding did not differ significantly by HIV status. Among participants with HHV-8 detected, shedding rates did not differ significantly by group. Median per-participant viral loads among positive samples were lowest in HIV+/KS+ (3.1 log_10_ copies/mL) and HIV-/KS+ participants (3.3 log_10_ copies/mL) relative to HIV+/KS-(3.8 log_10_ copies/mL) and HIV-/KS-participants (4.0 log_10_ copies/mL). All groups had participants with low-viral load intermittent shedding and those with high-viral load persistent shedding. Within each group, individual HHV-8 shedding rate positively correlated with median HHV-8 log_10_ copies/mL, and episode duration positively correlated with peak viral load.

**Conclusions:** Oral HHV-8 shedding is highly heterogeneous across Ugandan adults with and without KS and HIV. Persistent shedding is associated with higher median viral loads regardless of HIV and KS status.

## Introduction

Human herpes virus-8 (HHV-8) is the etiologic agent of Kaposi sarcoma (KS), a common cancer in sub-Saharan Africa (SSA) where HHV-8 seroprevalence is high. Several studies over the last two decades have shown that oral HHV-8 shedding is common in populations in SSA, ranging from 15-48% of individuals tested.[1–11] HHV-8 shedding in saliva is more frequent and occurs at a higher quantity, than in the genital tract and plasma, and behavior with exposure to saliva have been associated with HHV-8 infection.[12] Taken together, oral exposure may be an important driver of transmission.[13, 14] Oral shedding also predicts subsequent HHV-8 viremia, which may lead to infection of vascular or lymphatic endothelial cells and ultimately the development of KS.[14]

There is wide heterogeneity in HHV-8 shedding phenotypes among people infected with HHV-8, with 66% of shedding rate variance attributable to differences between individuals.[8] HHV-8 oral shedding frequency correlates with high median viral load; [14, 15]some individuals have brief, episodic, low viral load, oral shedding while others demonstrate persistent high viral loads.[8] HHV-8 shedding kinetics are at least partially modulated by the presence of KS and the degree of immunosuppression due to HIV co-infection. A previous study in Uganda demonstrated that oral HHV-8 shedding and viremia occur at higher frequency in adults with KS (KS+) versus no KS (KS–), with less substantial increases in HHV-8 shedding rate attributable to HIV infection.[14]

The mucosal shedding patterns of other human herpes viruses have been characterized in greater detail than for HHV-8 and represent the interactions between viral replication and spread and local immune responses.[16] Whereas genital herpes simplex virus 2 (HSV-2) infection is characterized by frequent bursts of replication which are rapidly eliminated over hours,[17–19] cytomegalovirus (CMV) has considerably slower kinetics with oral viral expansion and contraction phases lasting for weeks and months.[20, 21] Epstein-Barr virus (EBV) oral shedding varies substantially between individuals with evidence of limited episodic and high viral load persistence.[22, 23] For HSV-2 and EBV, the degree of HIV-1-related CD4+ T cell depletion inversely correlates with shedding rate, though only some variance in individual shedding rates can be attributed to an individual’s CD4+ T cell count.[22, 24] In this study, we aimed to evaluate individual-level HHV-8 viral kinetics to compare oral HHV-8 shedding rates, quantity, and episode characteristics among participants classified by HIV and KS status.

## Methods

### Study Design

We evaluated data from an observational, prospective cohort study of Ugandan adults (age ≥18 years) who were enrolled between October 2007 and May 2010. Individuals from four groups were invited to participate: HIV-1 seropositive individuals without KS (HIV+/KS-, Arm A); HIV-1 seronegative individuals without KS (HIV-/KS-, Arm B); HIV-1 seropositive individuals with KS (HIV+/KS+, Arm C); and HIV-1 seronegative individuals with KS (HIV-/KS+, Arm D). KS-negative participants (Arms A and B) were recruited from Voluntary Counseling and Testing Centers and HIV care centers in Kampala, Uganda. HIV+/KS-participants who reported ART use at the time of enrollment were excluded. HIV+/KS+ participants were eligible despite ART use. At the time of the original study, ART was initiated based on CD4 level, and universal test and treat policies were not yet established. Participants with a new diagnosis of KS (Arms C and D) were recruited from the Infectious Disease Institute and oncology clinics and wards at the Uganda Cancer Institute in Kampala, Uganda.

The enrollment visit included a rapid HIV test to determine HIV status, a physical exam to determine KS status, and for those who were KS positive, a skin biopsy of a suspected KS lesion or histologic confirmation. No HHV-8 serology tests were performed. Participants from all arms were followed for at least 4 weeks, with one “session” of data collection consisting of 28 days of daily home oral swab collection and weekly clinic visits that included physical exams, oral and anogenital mucosal swab collection, and plasma sample collection. HIV-positive individuals also had CD4 T-cell counts, and HIV viral loads measured at the first follow-up visit. This monthly cycle of sample collection was repeated every three months for Arm A (HIV+/KS-) participants (up to two years) and Arm C (HIV+/KS+) participants (up to one year).

This study was approved by the Makerere University Research and Ethics Committee, the Fred Hutch Cancer Center Institutional Review Board, and the University of Washington IRB.

### Definitions and Laboratory Methods

An HHV-8 shedding episode was defined by a string of consecutive positive days, allowing for one or more instances of a single missing or negative day within the string of positives. Episodes with well-defined durations were those with episode starts preceded by 2 consecutive negatives and episode stops followed by 2 consecutive negatives.

Oral swab samples were evaluated for HHV-8 DNA by quantitative, real-time polymerase chain reaction (PCR) at the UCI-Fred Hutch Cancer Centre Laboratory in Kampala, Uganda as described previously.[13, 15] Samples with >150 copies per mL of HHV-8 DNA were considered positive.[25] CD4 T-cell counts were measured by flow cytometry. HIV-1 RNA levels were measured with real-time RT PCR; levels <400 copies/mL were considered undetectable.

### Statistical Analysis

To ensure comparable sampling periods, data from the first 28-day session was used for comparisons between study arms. HHV-8 shedding rates were defined by the total number of days with HHV-8 detected divided by the total days with swabs collected and were computed for each participant (per-participant rates) as well as among all days contributed by participants in a study arm (day-level rates). HHV-8 quantity was computed among days with HHV-8 detected and measured in log_10_ copies/mL. We used negative binomial hurdle models,[26, 27] with an offset to account for differential follow-up time, to compare shedding rates by arm. These models accommodate excess zeros by including two components: 1) associations with zero shedding, where model estimates are presented as odds ratios (OR) with 95% confidence intervals (CI), and 2) associations with the rate of shedding among those with at least one day with HHV-8 detected, where model estimates are presented as incidence rate ratios (IRR) with 95% CIs. Shedding quantity on the log_10_ scale was compared between arms using generalized estimating equations (GEE) with the normal distribution, using methods to address informative cluster size,[28] as detailed in the supplement; model estimates were presented as mean log_10_ differences with 95% CIs. Arm B (HIV-/KS-) was considered the reference category in all models. We reported unadjusted estimates as well as model estimates adjusted for sex, age, alcohol use, lifetime number of sexual partners, and a variable with the following categories, to capture the extent of protected sexual contacts in the 3 months prior to enrollment: no partners in past 3 months, 100% condom use with sexual partners in past 3 months, <100% condom use with sexual partners in past 3 months. Sex and age were chosen as basic demographic adjustment variables; the remaining variables were chosen as these, or similar behavioral variables have shown significant associations with oral HHV-8 shedding rate in previous studies.[14]

We used descriptive statistics to characterize all HHV-8 episodes in each arm. We also characterized the episodes with the maximum observed duration for each participant; episode durations were compared by arm using survival methodology to treat those with episode duration not well-defined as censored observations, and maximum HHV-8 log_10_ copies/mL were compared using the Kruskal-Wallis test and subsequent pairwise tests that accounted for multiple comparisons. SAS, version 9.4 (SAS Institute, Cary, North Carolina) was used for all analyses.

## Results

### Cohort description

A total of 295 participants were enrolled: 125 were HIV+/KS-, 76 were HIV-/KS-, 76 were HIV+/KS+, and 18 were HIV-/KS+. Participant characteristics at enrollment are shown for the entire cohort and by study arm in **Table 1**. Overall, 134 (45%) were male and the median age was 35 years (range 18-71 years). Those with KS had a higher proportion of males than those without KS, and the HIV-/KS+ group had a higher median age than the other groups. Among HIV seropositive participants, CD4 counts were notably lower among individuals with KS (median 161 cells/μL, interquartile range [IQR] 48-273) than those without KS (median 411 cells/μL, IQR 277-598). The two HIV seropositive groups had similar proportions of participants with detectable HIV viral load (68% for each), but among participants with detectable HIV RNA, the HIV+/KS+ group had higher HIV viral load (median 5.0 log_10_ copies/mL, IQR 4.3-5.5) than the HIV+/KS-group (median 4.5 log_10_ copies/mL, IQR 4.2-5.0). Thirty-five of 76 (46%) participants in the HIV+/KS+ group reported previous ART; however, no previous ART use was reported by the HIV+/KS-group participants.

**Table 1.** Baseline Characteristics of Study Participants^a^.

| Characteristic | Total | HIV+/KS- | HIV-/KS- | HIV+/KS+ | HIV-/KS+ |
| --- | --- | --- | --- | --- | --- |
| Number of participants, n | 295 | 125 | 76 | 76 | 18 |
| **Demographic Factors** |  |  |  |  |  |
| Male | 134 (45%) | 37 (30%) | 29 (38%) | 50 (66%) | 18 (100%) |
| Age in years, median (range) | 35 (18 - 71) | 35 (18 - 64) | 34 (18 - 64) | 36 (19 - 58) | 44 (24 - 71) |
| **Clinical Factors** |  |  |  |  |  |
| BMI less than 18.5 | 38 (13%) | 13 (10%) | 9 (12%) | 13 (17%) | 3 (17%) |
| CD4 count<sup>b</sup>, median (IQR) | 305<br>(183 - 501) | 411<br>(277 - 598) |  | 161<br>(48 - 273) |  |
| Detectable HIV plasma RNA<sup>b</sup> | 134 (68%) | 84 (68%) |  | 50 (68%) |  |
| Log10 c/ml HIV plasma RNA<sup>b</sup>, median (IQR) | 4.6 (4.2 - 5.1) | 4.5 (4.2 - 5.0) |  | 5.0 (4.3 - 5.5) |  |
| KS any oral lesion<sup>c</sup> | 25 (27%) |  |  | 25 (33%) | 0 (0%) |
| KS Tumor Stage T1<sup>c</sup> | 82 (87%) |  |  | 66 (87%) | 16 (89%) |
| Received ART Prior to Session 1<sup>d</sup> | 36 (18%) | 0 (0%) |  | 35 (46%) |  |
| Received ART Concurrently with Session 1<sup>d</sup> | 48 (24%) | 8 (6%) |  | 40 (53%) |  |
| Received Chemotherapy Prior to Session 1<sup>d</sup> | 8 (9%) |  |  | 8 (11%) | 0 (0%) |
| Received Chemotherapy<br>Concurrently with Session 1<sup>d</sup> | 29 (31%) |  |  | 24 (32%) | 5 (28%) |
| **Behavioral Factors** |  |  |  |  |  |
| Drink alcohol | 111 (38%) | 47 (38%) | 34 (45%) | 23 (30%) | 7 (39%) |
| Lifetime sex partners, median<br>(IQR) | 5 (3 - 10) | 5 (3 - 10) | 4 (2 - 6) | 6 (5 - 13) | 5 (4 - 8) |
| Any sex partners in past 3<br>months | 160 (55%) | 67 (54%) | 53 (71%) | 27 (36%) | 13 (72%) |
| Condom use last 3 months |  |  |  |  |  |
| All the time | 55 (34%) | 31 (46%) | 12 (23%) | 12 (44%) | 0 (0%) |
| More than half the time | 21 (13%) | 16 (24%) | 4 (8%) | 1 (4%) | 0 (0%) |
| Less than half the time | 18 (11%) | 3 (4%) | 9 (17%) | 5 (19%) | 1 (8%) |
| Never | 66 (41%) | 17 (25%) | 28 (53%) | 9 (33%) | 12 (92%) |
| <sup>a</sup>Numbers may not add to totals due to missing data. Numbers shown as n(%) unless otherwise specified. |  |  |  |  |  |
| <sup>b</sup>Among HIV-positive participants only. HIV plasma RNA log10 c/ml only summarized among samples with detectable HIV plasma RNA. Maximum value (750000 c/ml) used for samples above maximum limit of detection. |  |  |  |  |  |
| <sup>c</sup>Among KS-positive participants only. |  |  |  |  |  |
| <sup>d</sup>Session 1 refers to the first period of 28 days of daily home oral swab collection |  |  |  |  |  |

### HHV-8 Shedding Rates and Quantity by Study Arm

During session one, oral swabs were collected for a median of 27-28 days for all study arms (**Table 2**). Twenty-six percent of KS-negative participants had HHV-8 detected in at least one oral swab, whereas among KS+ participants, 50% of HIV-/KS+ participants and 74% of HIV+/KS+ participants had HHV-8 detected in at least one oral swab. Correspondingly, participant-level shedding rates were higher among those with KS than those without KS (**Figure 1**); among those with at least one day with HHV-8 detected, the median shedding rate was 82% for the HIV-/KS+ group, 40% for the HIV+/KS+ group, 45% for the HIV-/KS-group and 51% for the HIV+/KS-group **(Figure 2b)**. Negative binomial hurdle models demonstrated that the higher shedding rates among KS+ participants were due to greater odds of any shedding; among those with HHV-8 detected in oral swabs on at least one day, there were no significant differences in frequency of HHV-8 shedding among study arms **(Supplemental Figure 1)**. The models also showed that participants with KS had greater odds of any shedding regardless of HIV status, whereas no significant differences in odds of shedding between HIV-positive and HIV-negative participants were detected when comparing participants with the same KS status **(Supplemental Figure 1)**. HHV-8 quantity among days with HHV-8 detected was lower among participants with KS and HIV, compared to those without KS or HIV (**Figure 2a-2b, Supplemental Figure 2**).

**Figure 1.**
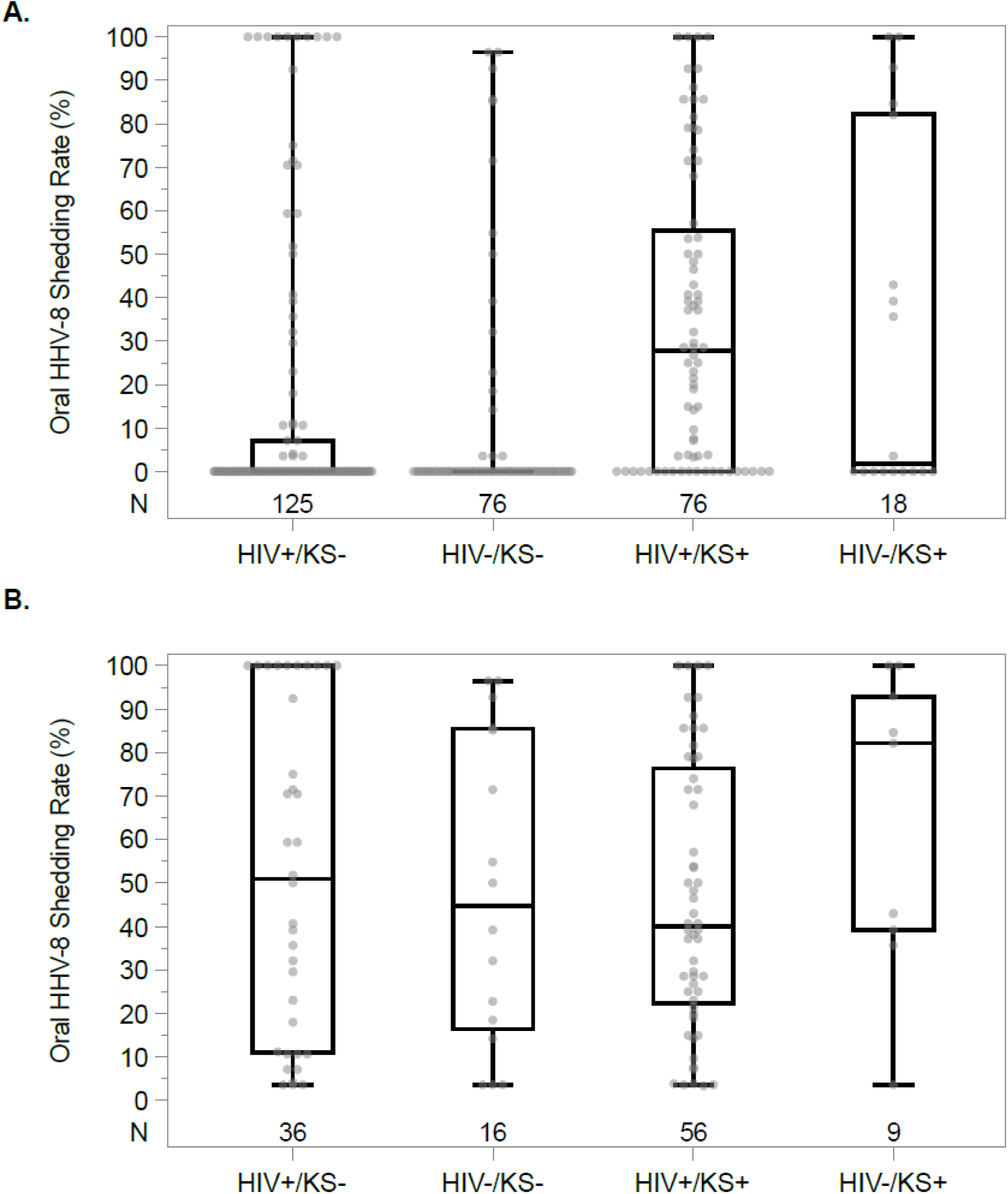
Distribution of participant-level oral HHV-8 shedding rate in session one by study arm. **A.** Among all participants. **B.** Among participants with HHV-8 detected orally on at least one day. Each data point represents a participant. Boxes represent the interquartile range, horizontal lines within boxes represent the medians, and whiskers extend to the minimum and maximum values. The absence of a box indicates that the 25^th^ and 75^th^ percentiles were both 0%. Numbers shown above the x-axis represent the number of participants contributing data in each arm.

**Figure 2.**
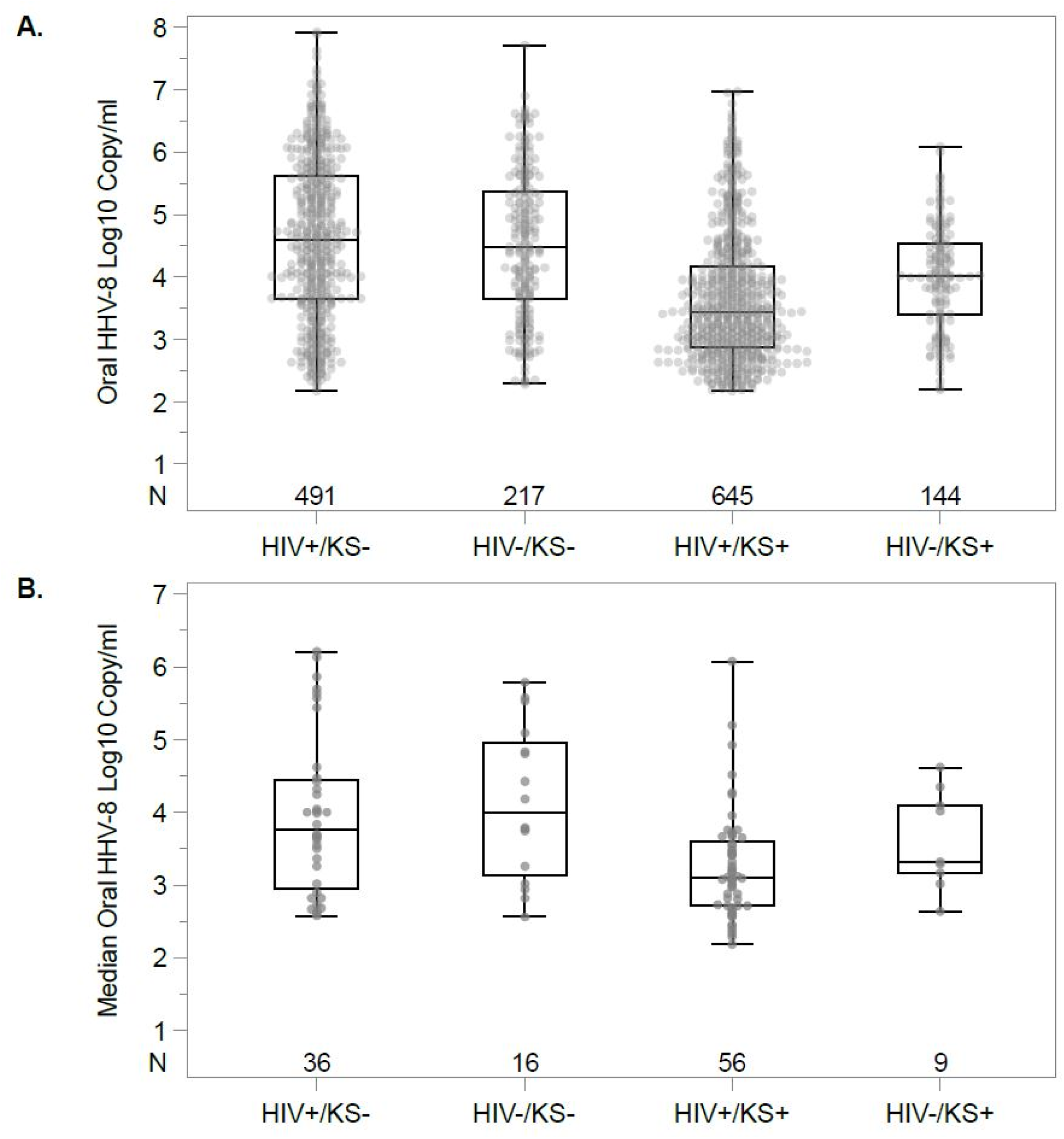
Distribution of oral HHV-8 log_10_ copies/mL by study arm. **A.** Among all days with HHV-8 detected in session one. Each data point represents a sample with HHV-8 detected; participants may contribute multiple data points. **B.** Median value per participant among days with HHV-8 detected in session one. Boxes represent the interquartile range, horizontal lines within boxes represent the medians, and whiskers extend to the minimum and maximum values. Sample size for each arm is shown just above the x-axis.

**Table 2.**
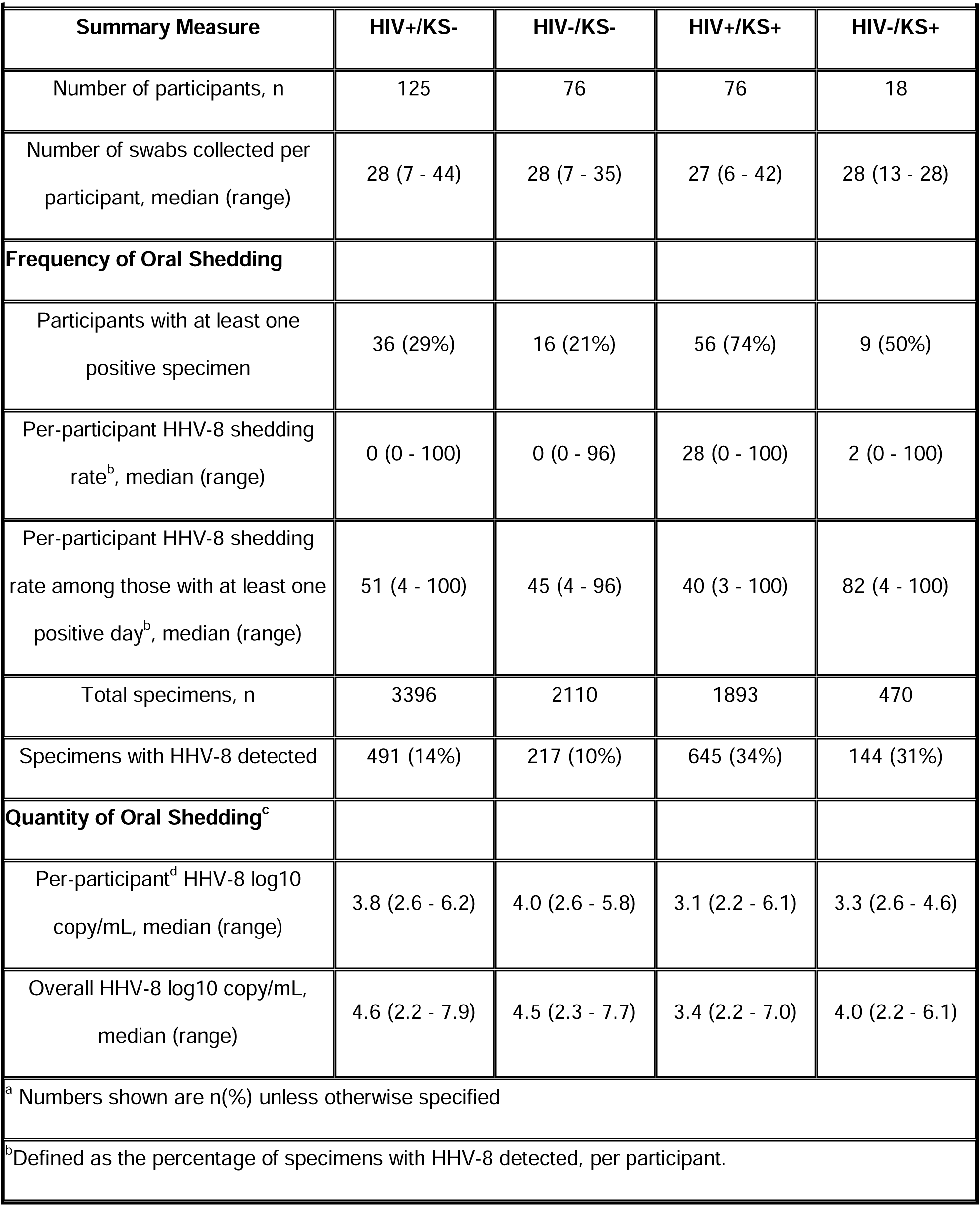
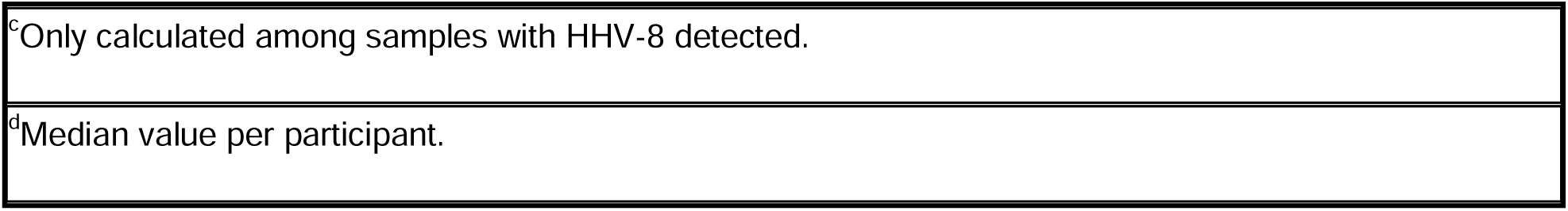
HHV-8 Frequency and Quantity Summaries^a^.

| Summary Measure | HIV+/KS- | HIV-/KS- | HIV+/KS+ | HIV-/KS+ |
| --- | --- | --- | --- | --- |
| Number of participants, n | 125 | 76 | 76 | 18 |
| Number of swabs collected per participant, median (range) | 28 (7 - 44) | 28 (7 - 35) | 27 (6 - 42) | 28 (13 - 28) |
| **Frequency of Oral Shedding** |  |  |  |  |
| Participants with at least one positive specimen | 36 (29%) | 16 (21%) | 56 (74%) | 9 (50%) |
| Per-participant HHV-8 shedding rate<sup>b</sup>, median (range) | 0 (0 - 100) | 0 (0 - 96) | 28 (0 - 100) | 2 (0 - 100) |
| Per-participant HHV-8 shedding rate among those with at least one positive day<sup>b</sup>, median (range) | 51 (4 - 100) | 45 (4 - 96) | 40 (3 - 100) | 82 (4 - 100) |
| Total specimens, n | 3396 | 2110 | 1893 | 470 |
| Specimens with HHV-8 detected | 491 (14%) | 217 (10%) | 645 (34%) | 144 (31%) |
| **Quantity of Oral Shedding<sup>c</sup>** |  |  |  |  |
| Per-participant<sup>d</sup> HHV-8 log10 copy/mL, median (range) | 3.8 (2.6 - 6.2) | 4.0 (2.6 - 5.8) | 3.1 (2.2 - 6.1) | 3.3 (2.6 - 4.6) |
| Overall HHV-8 log10 copy/mL, median (range) | 4.6 (2.2 - 7.9) | 4.5 (2.3 - 7.7) | 3.4 (2.2 - 7.0) | 4.0 (2.2 - 6.1) |

<sup>a</sup> Numbers shown are n(%) unless otherwise specified
<sup>b</sup> Defined as the percentage of specimens with HHV-8 detected, per participant.
<sup>c</sup>Only calculated among samples with HHV-8 detected.
<sup>d</sup>Median value per participant.

### HHV-8 Shedding Episode Characteristics by Study Arm, Among Those with HHV-8 Detected

Participants had a total of 230 episodes of oral HHV-8 shedding, with a median of 1 episode per participant for all study arms except for the HIV+/KS+ arm, where the median was 2 episodes per participant (**Table 3**). Fewer than half of the episodes had a well-defined start and stop; thus, durations computed among observed data were likely underestimates. Using the observed data, the median durations among all episodes ranged from 2 days in the HIV+/KS+ arm, 5 days in the HIV+/KS-and HIV-/KS-arms, and 10 days in the HIV-/KS+ arm. One-day episodes were common and comprised anywhere from 23% of episodes among HIV-/KS+ participants to 40% of episodes among HIV+/KS+ participants. Median peak HHV-8 viral load per episode was highest in the HIV-/KS+ group (4.5 log_10_ copies/mL) and lowest in the HIV+/KS+ group (3.2 log_10_ copies/mL).

**Table 3.** HHV-8 Episode Characteristics of Study Participants^a^.

| **Episode Summary** | **Total** | **HIV+/KS-** | **HIV-/KS-** | **HIV+/KS+** | **HIV-/KS+** |
| --- | --- | --- | --- | --- | --- |
| Total number of episodes<sup>a</sup>, n | 230 | 59 | 29 | 129 | 13 |
| Number of episodes per participant, median (range) | 2 (1 - 6) | 1 (1 - 6) | 1 (1 - 5) | 2 (1 - 6) | 1 (1 - 2) |
| **Episode-Level Summaries** |  |  |  |  |  |
| Episodes with well-defined duration<sup>b</sup> | 102 (44%) | 23 (39%) | 13 (45%) | 63 (49%) | 3 (23%) |
| Episode duration in days (naive<sup>c</sup>), median (range) | 3 (1 - 35) | 5 (1 - 29) | 5 (1 - 28) | 2 (1 - 35) | 10 (1 - 28) |
| Episodes of 1-day duration | 83 (36%) | 20 (34%) | 8 (28%) | 52 (40%) | 3 (23%) |
| Minimum log10 copy/mL per episode, median (IQR) | 2.6 (0.0 - 5.8) | 2.7 (0.0 - 5.8) | 2.8 (0.0 - 4.7) | 2.5 (0.0 - 5.1) | 2.7 (0.0 - 3.0) |
| Maximum log10 copy/mL per episode, median (IQR) | 3.7 (2.2 - 7.9) | 4.1 (2.3 - 7.9) | 4.1 (2.3 - 7.7) | 3.2 (2.2 - 7.0) | 4.5 (2.8 - 6.1) |
| Median log10 copy/ml per episode, median (IQR) | 3.1 (2.2 - 6.4) | 3.4 (2.3 - 6.4) | 3.6 (2.3 - 5.7) | 3.0 (2.2 - 6.1) | 3.4 (2.6 - 4.6) |
| **Participant-Level Summaries, Using Episode with Maximum Observed Duration per Participant** |  |  |  |  |  |
| Number of participants | 117 | 36 | 16 | 56 | 9 |
| Episodes with well-defined duration<sup>b</sup> | 36 (31%) | 9 (25%) | 4 (25%) | 22 (39%) | 1 (11%) |
| Episode duration in days (naive<sup>c</sup>), median (range) | 9 (1 - 35) | 10 (1 - 29) | 8 (1 - 28) | 8 (1 - 35) | 13 (1 - 28) |
| Episodes of 1-day duration | 21 (18%) | 7 (19%) | 3 (19%) | 10 (18%) | 1 (11%) |
| Minimum log10 copy/mL per episode, median (IQR) | 2.5 (0.0 - 5.8) | 2.6 (0.0 - 5.8) | 1.3 (0.0 - 4.4) | 1.1 (0.0 - 4.9) | 2.7 (0.0 - 3.0) |
| Maximum log10 copy/mL per episode, median (IQR) | 4.5 (2.2 - 7.9) | 4.8 (2.6 - 7.9) | 5.0 (2.6 - 7.7) | 4.0 (2.2 - 7.0) | 4.8 (3.0 - 6.1) |
| Median log10 copy/mL per episode, median (IQR) | 3.4 (2.2 - 6.4) | 3.7 (2.5 - 6.4) | 3.7 (2.3 - 5.7) | 3.2 (2.2 - 6.1) | 3.5 (2.6 - 4.6) |
| <sup>a</sup>Episodes defined by consecutive positive days, allowing one or more instances of a single negative or missing value within the string of positives. Numbers shown are n(%) unless otherwise specified. |  |  |  |  |  |
| <sup>b</sup>Well-defined duration is defined by episode start preceded by 2 consecutive negatives and episode end followed by 2 consecutive negatives. |  |  |  |  |  |
| <sup>c</sup>Including episodes without well-defined duration, using observed durations. These are likely underestimates |  |  |  |  |  |

Selecting the episode with the maximum observed duration for each participant, we performed some additional comparisons among study arms. First, we grouped participants into five categories (maximum episode duration 1 day, 2-5 days, 6-10 days, >10 days with shedding rate <100%, and shedding rate=100%) based on observed episode duration and shedding rate. Individual shedding patterns for participants within each category are shown in **Supplemental Figure 3**. Over the 28-day sampling period, we observed both episodic and continuous HHV-8 shedding patterns. All study arms had at least one participant with a maximum duration episode of only one day. Episodes lasting more than 10 days were more common among the HIV-/KS-arm, although unlike the other study arms, there were no participants in that arm with continuous shedding during the sampling period. Next, we used survival methodology to compare episode duration among study arms, while treating episode durations that were not well-defined as censored observations; there was no significant difference in episode duration among the 4 study arms (p=0.26, log-rank test). Maximum HHV-8 copy number among these episodes of maximum duration per participant appeared lowest among the HIV+/KS+ group (Kruskal-Wallis p=0.054 for differences among the 4 arms; individual pairwise comparisons accounting for multiple comparisons not statistically significant).

### Relationship Between Shedding Rate, Quantity, and Episode Characteristics, Among Those with HHV-8 Detected

Among participants with HHV-8 detected on at least one day, per-participant oral HHV-8 shedding rate correlated with median oral HHV-8 log_10_ copy/mL among positive days, for all groups (**Figure 3**). When selecting the longest episode per participant, peak HHV-8 log_10_ copy/mL correlated with duration such that brief, single, or recurrent 1-10-day episodes generally had lower peak viral loads than episodes lasting >10 days, for all study arms (**Supplemental Figure 3-4)**. All groups had episodes of both long and short durations, and episodes lasting <10 days with peak viral loads <4 log_10_ copy/mL were common among the 4 groups: HIV+/KS+ group (23/56 (41%) participants), HIV+/KS-: 12/36 (33%), HIV-/KS-: 5/16 (31%) and HIV-/KS+: 2/9 (22%).

**Figure 3.**
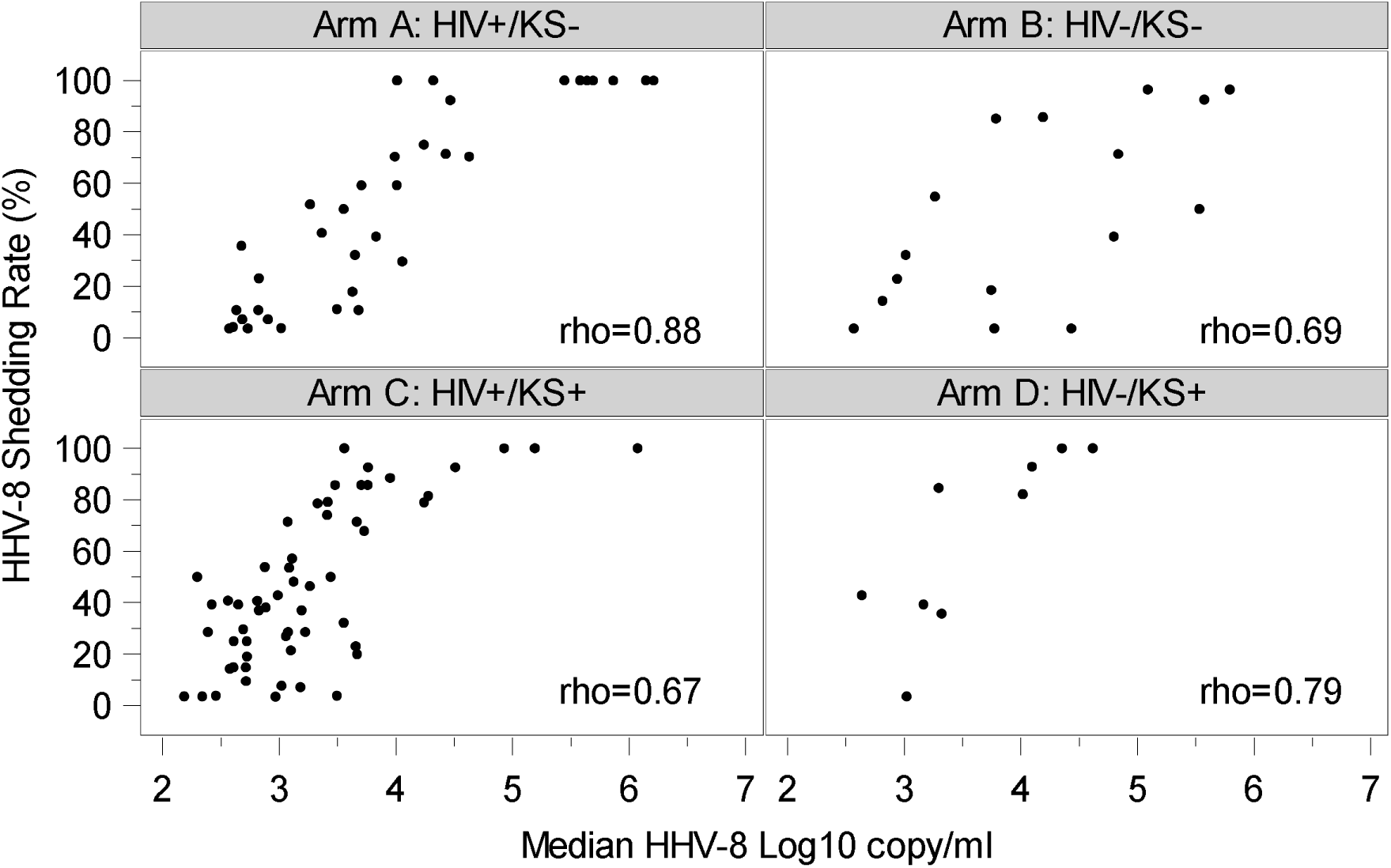
Oral HHV-8 shedding rate by median HHV-8 Log_10_ copy/mL, among positive days, for each study arm. Each data point represents a participant with at least one day with oral HHV-8 detected; Arm A (HIV+/KS-), n=36; Arm B (HIV-/KS-), n=16; Arm C (HIV+/KS+), n=56; Arm D (HIV-/KS+), n=9. Data have been jittered to allow viewing of overlapping points. Spearman correlation coefficients are shown for each arm; pairwise comparisons of correlations for each study arm did not show significant differences when adjusting for multiple comparisons.

**Figure 4.**
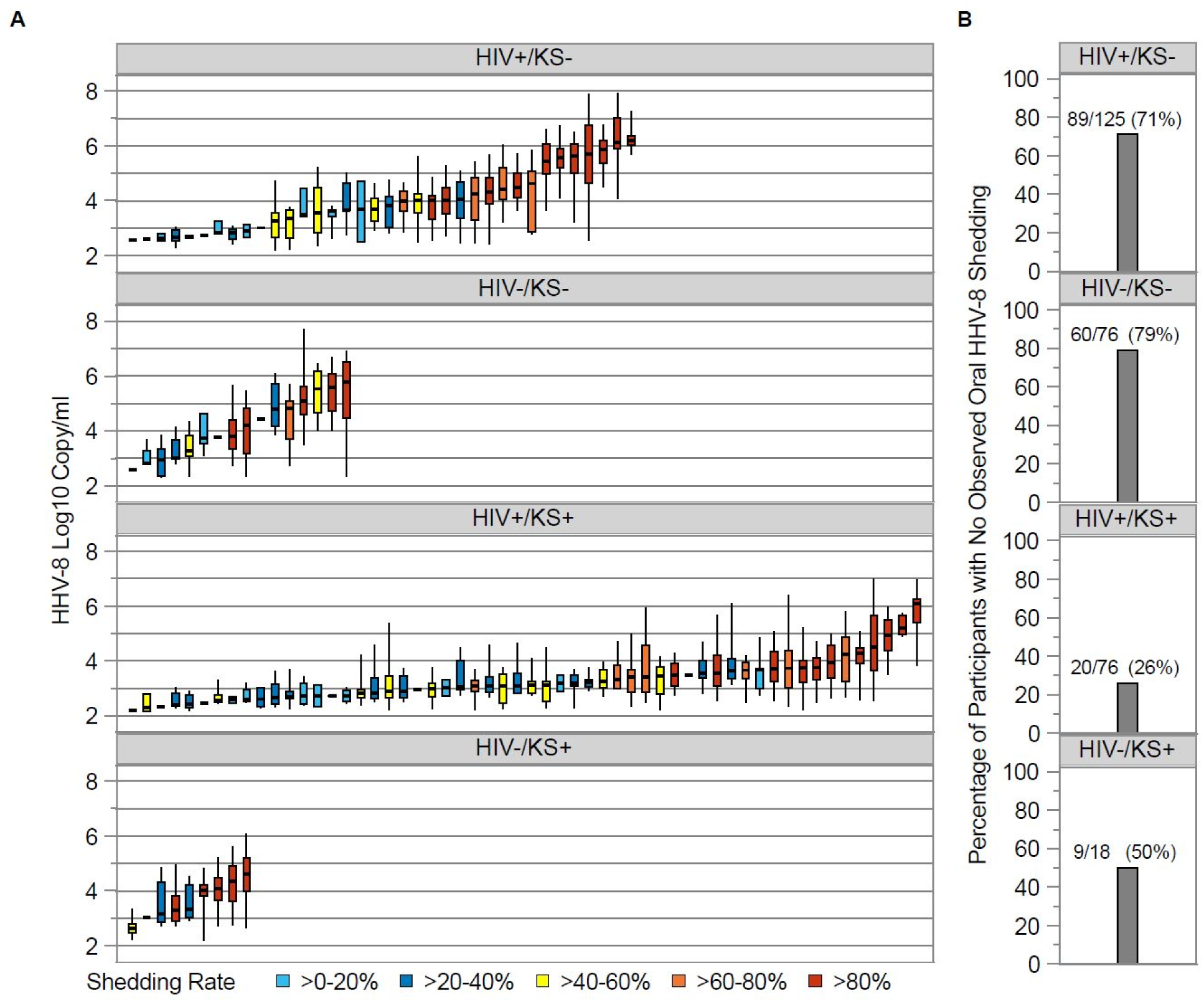
**A.** Distribution of oral HHV-8 viral loads for individual participants, using session 1 data only, by study arm. Each boxplot shows data from a participant; boxes represent interquartile range, horizontal lines within boxes represent medians and whiskers extend to minimum and maximum values. Participants with only a median bar and no box had too few positive samples to generate an interquartile range. Participants are sorted along the x-axis by median HHV-8 log10 copy/mL and boxes are color coded according to shedding rate among all session 1 samples. **B.** Percentage of participants with no observed oral HHV-8 during session 1, by study arm. Numbers atop the bars show the number of participants with no observed shedding over the total number of participants in each arm and computed percentage.

Among individual shedders in all 4 groups, high-frequency shedders typically had high median viral loads among positive samples whereas shedding rates < 60% were usually associated with lower median viral loads **(Figure 3-4)**. Considerable variability was noted in viral load within individuals as well with viral loads occasionally varying over several orders of magnitude **(Figure 4).**

### Within-person correlation in HHV-8 shedding over longer follow-up among HIV+/KS-persons

Among the 125 HIV+/KS-participants, 92 collected oral swabs in more than one defined time category from the start of session one and thus contributed data to evaluate shedding patterns over a longer period **(Supplemental Methods**). The 92 individuals contributed a median of five time categories each (range 2-6) over a median of 15.8 months of follow-up (range 3.6-25.4 months). The time categories were comprised of a median of 28 oral swabs collected (range 7-60). **Supplemental Figure 5** shows oral HHV-8 shedding rates by time category for these participants. Though some participants displayed high variability in shedding rates, the majority (71/92, 77%) demonstrated ≤30% absolute change in shedding rates, compared to the initial time category. Correlations between shedding rates in later time categories and the initial shedding rate were generally high (**Supplemental Figure 6**). However, model estimates adjusted for CD4 count and HIV viral load showed significantly lower shedding rates in days 210 to <330 compared to the initial shedding rate, which generally did not persist (**Supplemental Figure 7**).

Patterns in median oral HHV-8 copy/mL among 28 HIV+/KS-participants contributing >1 time category with at least one day with HHV-8 detected are shown in **Supplemental Figure 8**. Of the 28 participants, 13 (46%) had follow-up median copy number vary <1 log_10_ compared to the initial median copy number. Moderate correlations were observed between the initial and subsequent median log_10_ copy number (**Supplemental Figure 9**). Similar to shedding rates, adjusted model estimates showed HHV-8 quantity in days 210 to <330 was lower than the initial HHV-8 quantity, but this decrease did not persist (**Supplemental Figure 10**).

## Discussion

We describe the general pattern of HHV-8 oral shedding in a cohort of Ugandan adults. This cohort builds on past studies of HHV-8 oral shedding by specifically considering viral load and by differentiating episodic versus persistent shedding patterns at the individual level. We found that the presence of any shedding was more common in individuals with KS, regardless of HIV status, but all types of viral kinetics -- including no shedding, low viral load intermittent shedding, and persistent high viral load shedding -- were noted in all four groups. Oral shedding rate was similar between the four groups when only shedders were analyzed but viral load was lower by 0.9 log_10_ among HIV+ / KS+ individuals compared to HIV-/KS-individuals. This difference in viral loads was also reported in a previous, smaller study that measured HHV-8 shedding among Ugandan individuals grouped by KS and HIV status [13]; compared to that study, our larger study also found similar estimates of oral HHV-8 shedding rates within groups defined by presence of KS and HIV. We found a positive correlation between shedding rate and median viral load in all four groups. Overall, these results show deep variability in shedding kinetics among individuals, and these differences are only partially explained by KS or HIV co-infection. However, our study demonstrates that distinct shedding “phenotypes” of low versus high frequency and copy number oral HHV-8 shedding appear to persist over time, at least among HIV+/KS-individuals. Persons with persistent high-level HHV-8 shedding may be particularly important targets for interventions to reduce oral HHV-8 replication to prevent the development of KS in these individuals or to limit HHV-8 transmission to others.

These results raise several important unanswered questions that our study was not able to address. First, what drives the distinct observed shedding patterns other than HIV status and KS status? One key unmeasured variable in our study is the time of initial HHV-8 infection, as shedding frequency or quantity may be higher closer to primary infection. In Uganda, most persons are infected during adolescence, however, so we expect that most of our participants had primary HHV-8 infection for several years before study enrollment.[29, 30] Our longitudinal analyses of shedding rates and quantity also suggest that these outcomes are somewhat stable over time within individuals with some exceptions, raising the unanswered question of whether a shedding “set point” may be established soon after primary infection. Other potential key unmeasured variables in this current study are inoculum dose at the time of acquisition, viral strain, host immune parameters, recent re-infection, and other medical co-morbidities. In addition, we cannot be certain whether the KS-negative participants who did not demonstrate viral shedding were infected with HHV-8 given limitations in HHV-8 serologic testing, and the inclusion of some uninfected participants may partially explain the lower shedding rates in this group.

Our findings also suggest several avenues for future research. An important future focus will be on understanding how systemic and local immune responses influence viral shedding, including whether tissue-resident immune responses are responsible for the rapid elimination of local shedding in individuals whose infection manifests only as periodic low viral-load blips. Prospective cohorts that follow low and high shedders without KS over prolonged periods could help determine if persistent, high-copy number oral HHV-8 shedding is predictive of tumor development. If so, novel strategies directed at HHV-8 to prevent KS should be considered, including anti-HHV-8 antiviral therapies or HHV-8 vaccination. Oral valganciclovir lowers HHV-8 shedding rate though it remains unknown whether antiviral therapy has a role in preventing HHV-8 associated tumors.[14] Similarly, among individuals with KS, further research is needed to establish whether changes in oral viral shedding could serve as a biomarker for therapeutic response. Finally, another key area of future research is understanding the role of HHV-8 viral load in driving HHV-8 transmission. We have demonstrated in previous studies that household levels of cytomegalovirus and human herpes virus-6 are predictive of subsequent acquisition.[31, 32] To discern whether similar transmission dynamics are relevant for HHV-8 would require detailed household cohort studies that attempt to capture incident infection in young children.

In conclusion, we demonstrate substantial variability in HHV-8 oral shedding among Ugandan adults regardless of HIV and KS status, though individuals with KS are more likely to have detectable oral HHV-8 shedding. However, some individuals may have stable phenotypes of intermittent, low copy number shedding vs persistent, high copy number shedding. Understanding the biological mechanisms underpinning these differences could help guide the development of new biomarkers, strategies to reduce HHV-8 transmission, and ultimately the development of a new preventative or therapeutic HHV-8 vaccine or other immune-based therapeutic approaches.

## Data Availability

All data produced in the present work are contained in the manuscript

## Authorship contributions

E.M.K. assisted in the study design, performed the statistical analyses, interpreted results, and wrote the manuscript. J.T.S., C.C. and W.P. designed the study and provided funding for the work; W.P and J.T.S interpreted the results, provided clinical input, and wrote the manuscript. I.M., J.N., F.O., acquired data and reviewed the manuscript; D.S. interpreted results and reviewed the manuscript; C.C. and J.O. reviewed the manuscript.

## Potential conflicts of interest

JTS receives consulting fees from Pfizer for advising on SARS-CoV-2 therapeutics and Glaxo Smith Kline for advising on HSV vaccines. CC receives fees for consulting from Recordati Rare Disease, for sitting on the Scientific Advisory Board of Viracta Therapeutics, and receives research funding from ImmunityBio and Janssen. All other authors report no potential conflict of interest.

This information has been previously presented at the 25^th^ International Conference on Kaposi Sarcoma Herpesvirus and Related Agents. Dar es Salaam, Tanzania, June 19-23, 2023.

## Acknowledgements

We are grateful to the study participants and the study team, whose commitment made this study possible.

