## Supplemental Materials for "Highly heterogeneous human herpes virus-8 oral shedding kinetics among people with and without Kaposi sarcoma and HIV- co-infection"

**Supplemental Methods**

*Statistical Analysis*

For the comparison of HHV-8 shedding quantity among study arms, we needed to account for informative cluster size, where the number of observations per participant (number of days with HHV-8 detected) was associated with the outcome (HHV-8 log_10_ copies/mL). To do so, we used an independence working correlation with weights equal to the inverse of the number of observations per participant (1 / number of days with HHV-8 detected). By using this approach, our model estimates can be interpreted as the association between HIV/KS status and HHV-8 quantity for a typical positive day from a typical participant in each group.

To describe within-person patterns in HHV-8 shedding over a longer period among HIV seropositive participants, we focused on those without KS because a relatively low proportion of the HIV+/KS+ participants completed more than one follow-up session. To achieve reasonable sample sizes for longitudinal comparisons, we grouped sessions into the following time categories from the start of session one: 0 to <90 days, 90 to <210 days, 210 to <330 days, 330 to <450 days, 450 to <570 days, and 570 to 750 days. For both shedding rates and shedding quantity, we computed correlations between the initial time period and each follow-up time period. We also tested for differences over time using unadjusted models and models adjusted for HIV viral load and CD4 count measured at the start of each time category. For the shedding rate outcome, we used generalized linear mixed effects models with a negative binomial distribution, random intercept for each person, and empirical variance option to produce robust standard errors. For the HHV-8 quantity outcome, we used similar models but with the normal distribution. The initial time category was considered the reference category.

**Supplemental Results**

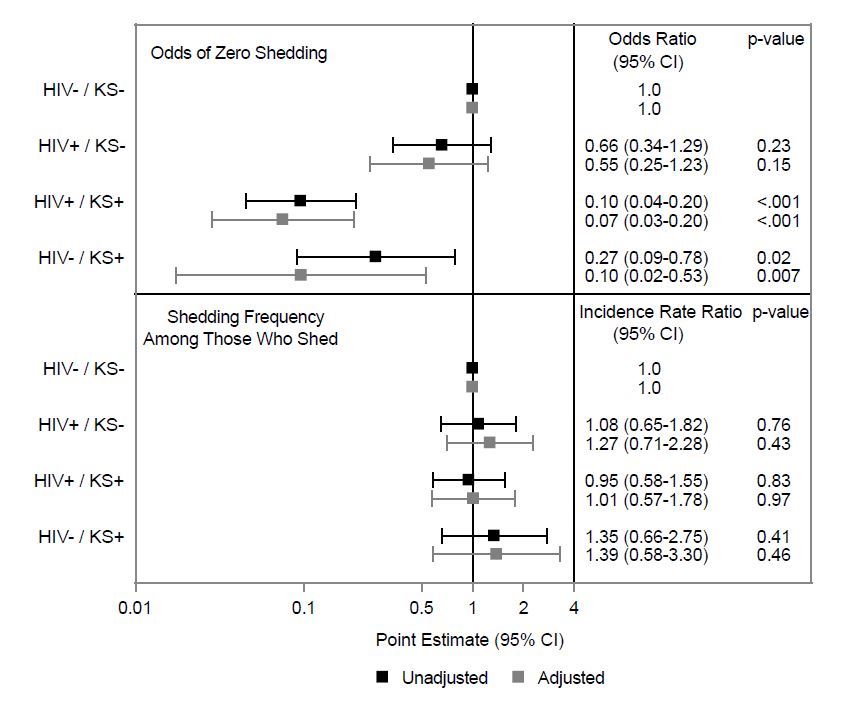

**Supplemental Figure 1.** Negative binomial hurdle model estimates comparing oral HHV-8 shedding rates among groups defined by HIV and KS status, using session 1 data. Models include two components: 1) associations with zero shedding where estimates are presented as odds ratios with 95% confidence intervals (CI) in the top portion of the figure and 2) associations with rate of shedding among those with at least one day with HHV-8 detected, where estimates are presented as incidence rate ratios with 95% CIs. These point estimates are depicted by the squares and 95% CIs are shown by the extending bars. Unadjusted estimates are shown in black and estimates adjusted for sex, age, alcohol use, number of lifetime sexual partners, and condom use with partners in the past 3 months are shown in grey. The vertical reference line at a point estimate of 1 represents no difference. Participants without HIV and without KS served as the reference category; estimates shown for the other 3 participant groups represent comparisons with the HIV- / KS- group.

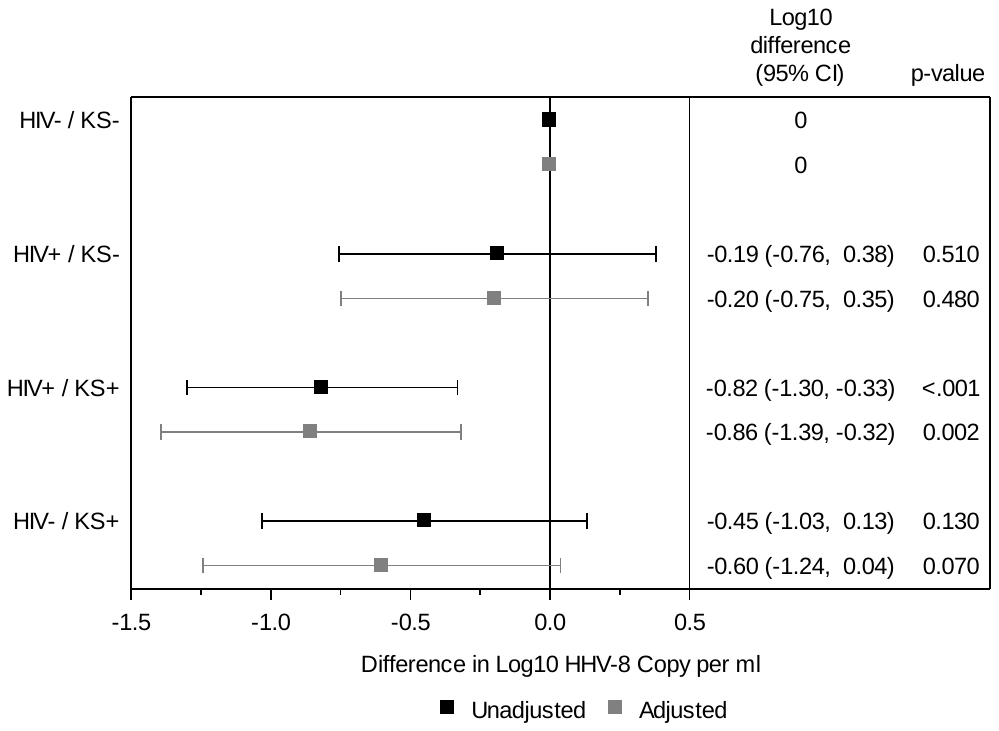

**Supplemental Figure 2.** Generalized estimating equations model estimates comparing oral HHV-8 shedding quantity among groups defined by HIV and KS status, using session 1 positive days. Models used the Normal distribution with identity link; estimates are presented as mean difference in log_10_ copy/ml, depicted by the squares, with 95% confidence intervals (CI), shown by the extending bars. Unadjusted estimates are shown in black and estimates adjusted for sex, age, alcohol use, number of lifetime sexual partners, and condom use with partners in the past 3 months are shown in grey. The vertical reference line at a difference of 0 represents no difference. Participants without HIV and without KS served as the reference category; estimates shown for the other 3 participant groups represent comparisons with the HIV- / KS- group.

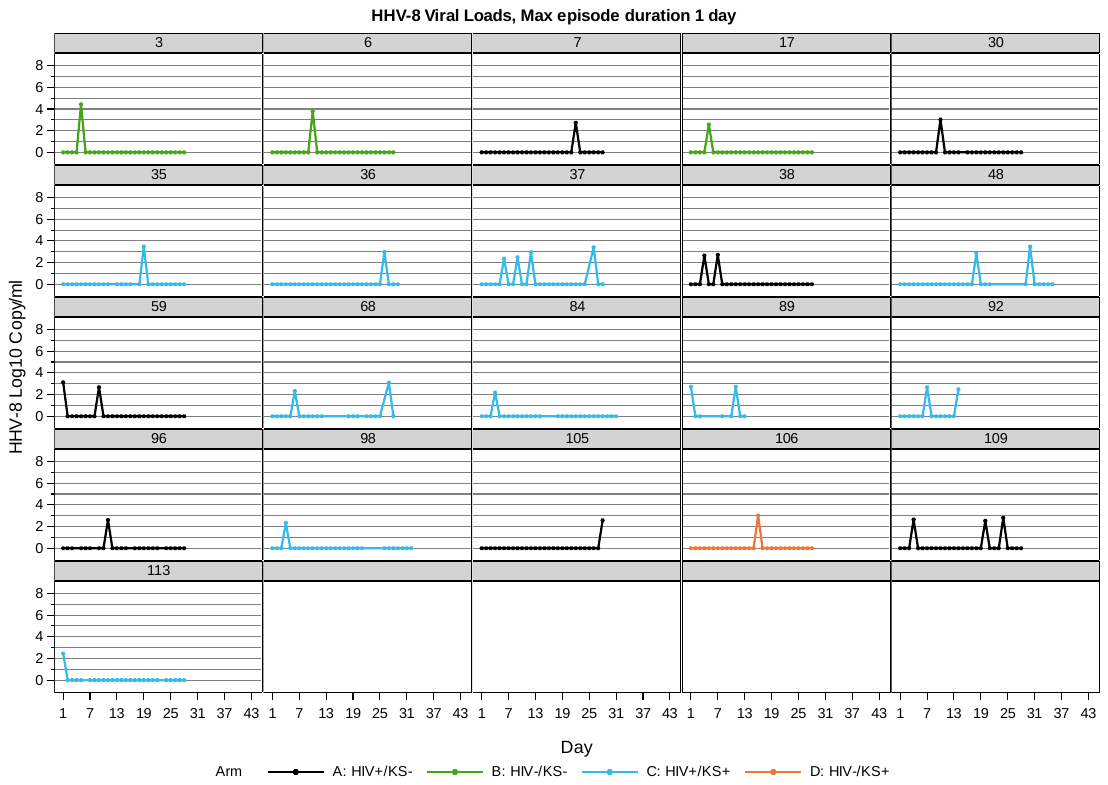

**Supplemental Figure 3a**. Individual participant plots of oral HHV-8 viral loads during session 1, among 21 participants with shedding rates < 100% and maximum episode duration of 1 day. Negative results are represented by a log_10_ value of 0. Line color indicates study arm, as specified in key. Of 36 total HIV+/KS- participants with HHV-8 detected, 7 (19%) met these criteria; of 16 HIV-/KS- participants with HHV-8 detected, 3 (19%) met these criteria; of 56 HIV+/KS+ participants with HHV-8 detected, 10 (18%) met these criteria; of 9 HIV-/KS+ participants with HHV-8 detected, 1 (11%) met these criteria.

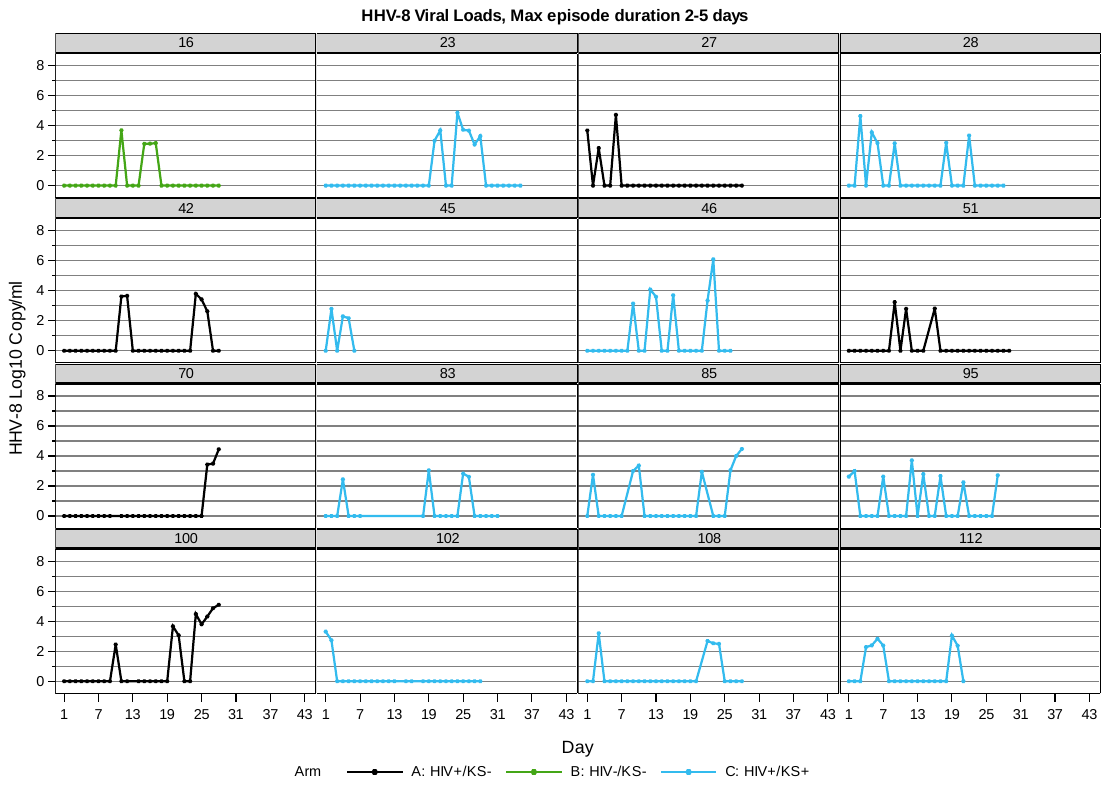

**Supplemental Figure 3b**. Individual participant plots of oral HHV-8 viral loads during session 1, among 16 participants with shedding rates < 100%, maximum episode duration 2-5 days. Negative results are represented by a log_10_ value of 0. Line color indicates study arm, as specified in key. Of 36 total HIV+/KS- participants with HHV-8 detected, 5 (14%) met these criteria; of 16 HIV-/KS- participants with HHV-8 detected, 1 (6%) met these criteria; of 56 HIV+/KS+ participants with HHV-8 detected, 10 (18%) met these criteria; of 9 HIV-/KS+ participants with HHV-8 detected, none met these criteria.

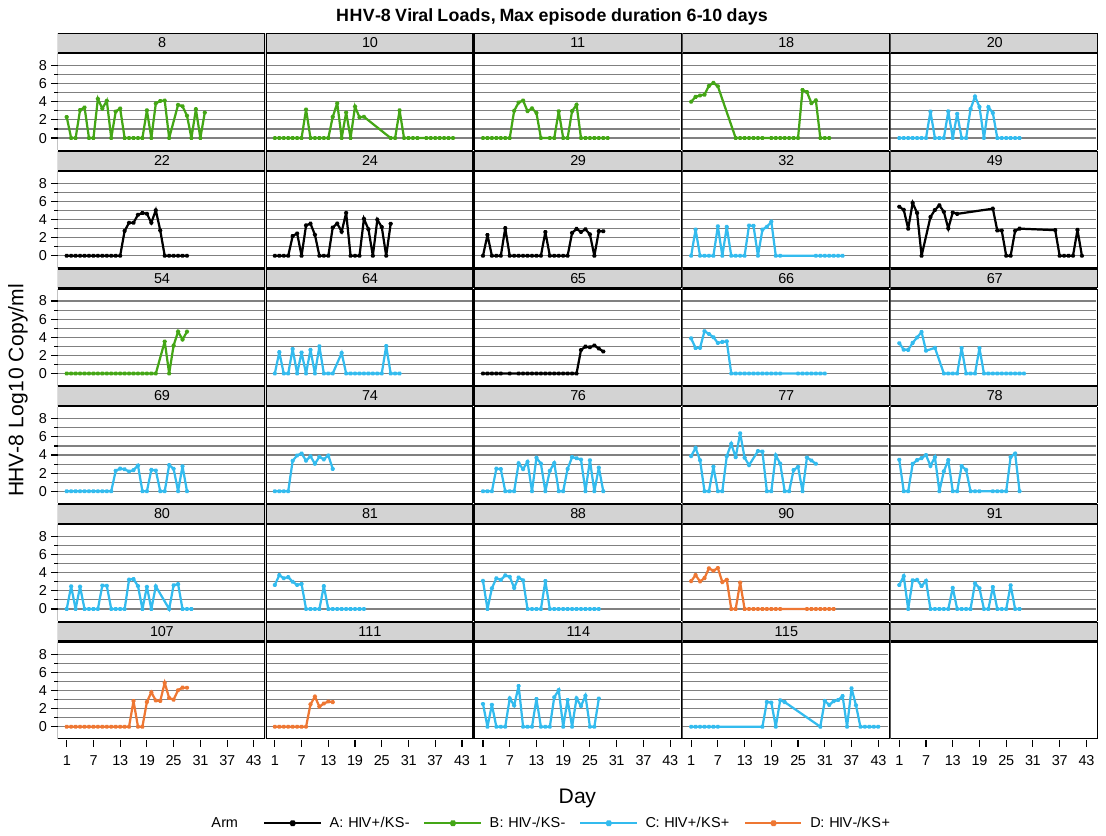

**Supplemental Figure 3c**. Individual participant plots of oral HHV-8 viral loads during session 1, among 29 participants with shedding rates < 100%, maximum episode duration 6-10 days. Negative results are represented by a log_10_ value of 0. Line color indicates study arm, as specified in key. Of 36 total HIV+/KS- participants with HHV-8 detected, 5 (14%) met these criteria; of 16 HIV-/KS- participants with HHV-8 detected, 5 (31%) met these criteria; of 56 HIV+/KS+ participants with HHV-8 detected, 16 (29%) met these criteria; of 9 HIV-/KS+ participants with HHV-8 detected, 3 (33%) met these criteria.

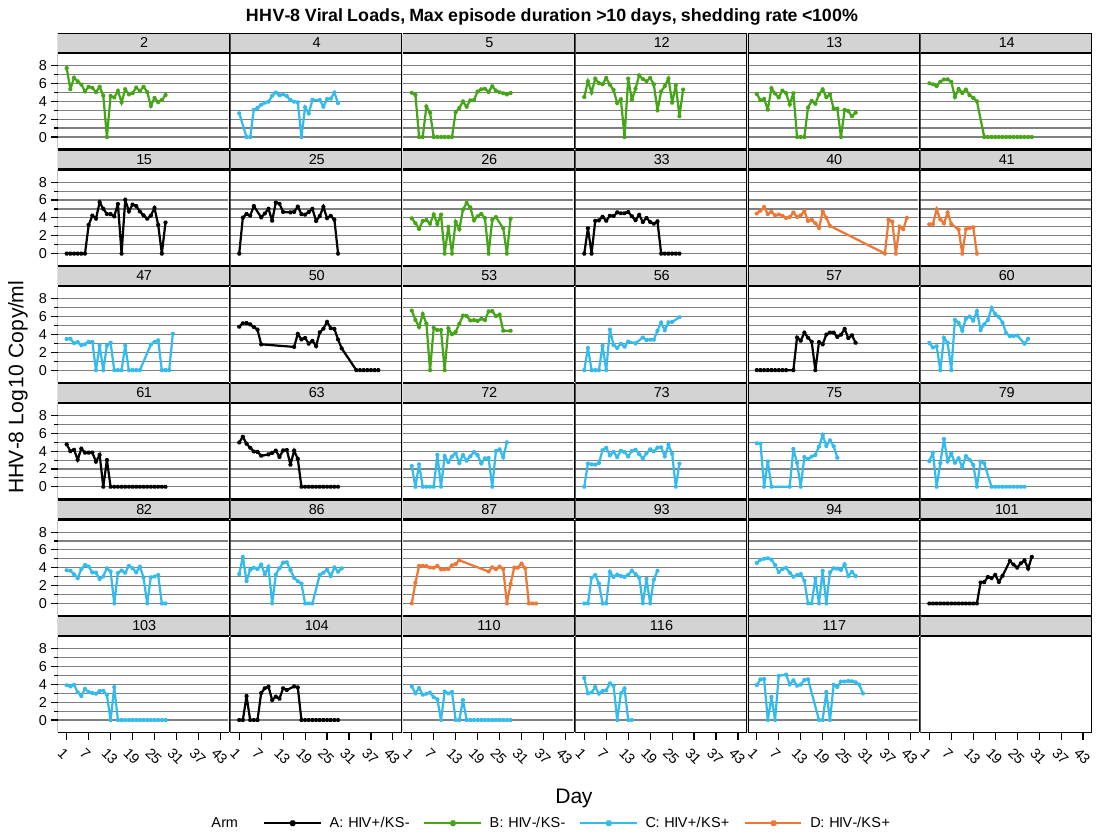

**Supplemental Figure 3d**. Individual participant plots of oral HHV-8 viral loads during session 1, among 35 participants with shedding rates < 100%, maximum episode duration >10 days. Negative results are represented by a log_10_ value of 0. Line color indicates study arm, as specified in key. Of 36 total HIV+/KS- participants with HHV-8 detected, 9 (25%) met these criteria; of 16 HIV-/KS- participants with HHV-8 detected, 7 (44%) met these criteria; of 56 HIV+/KS+ participants with HHV-8 detected, 16 (29%) met these criteria; of 9 HIV-/KS+ participants with HHV-8 detected, 3 (33%) met these criteria.

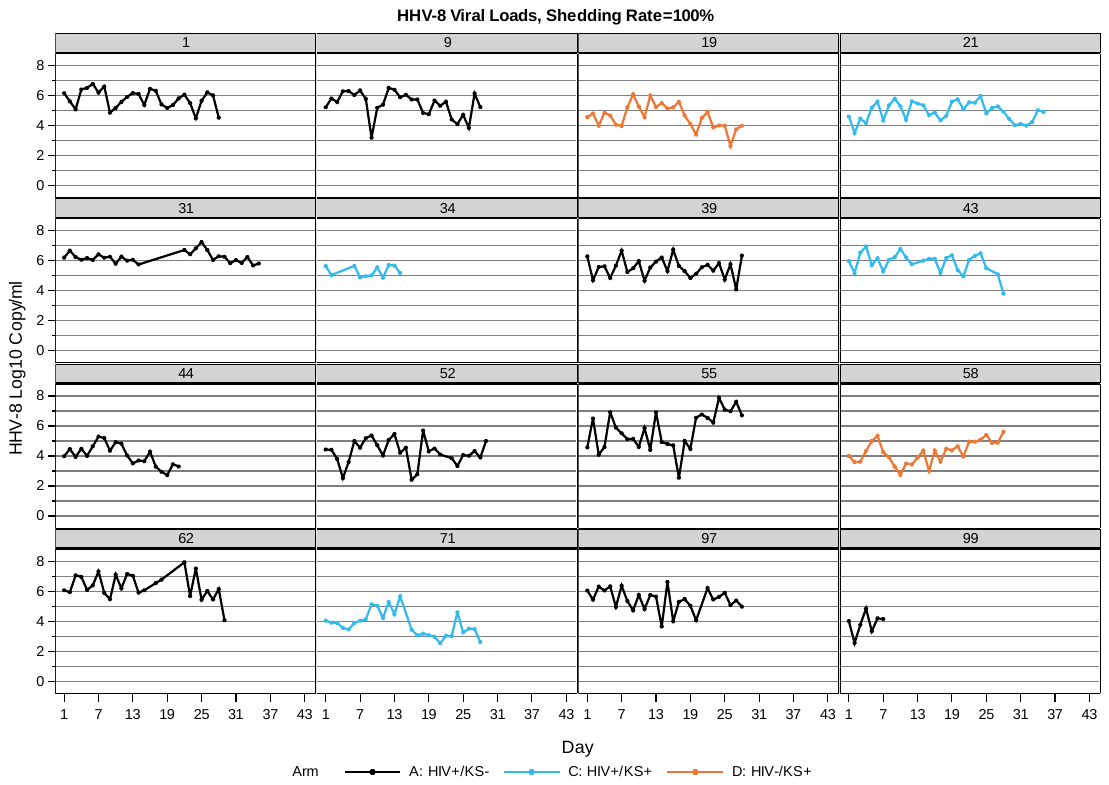

**Supplemental Figure 3e**. Individual participant plots of oral HHV-8 viral loads during session 1, among 16 participants with shedding rates of 100%. Negative results are represented by a log_10_ value of 0. Line color indicates study arm, as specified in key. Of 36 total HIV+/KS- participants with HHV-8 detected, 10 (28%) met these criteria; of 16 HIV-/KS- participants with HHV-8 detected, none met these criteria; of 56 HIV+/KS+ participants with HHV-8 detected, 4 (7%) met these criteria; of 9 HIV-/KS+ participants with HHV-8 detected, 2 (22%) met these criteria.

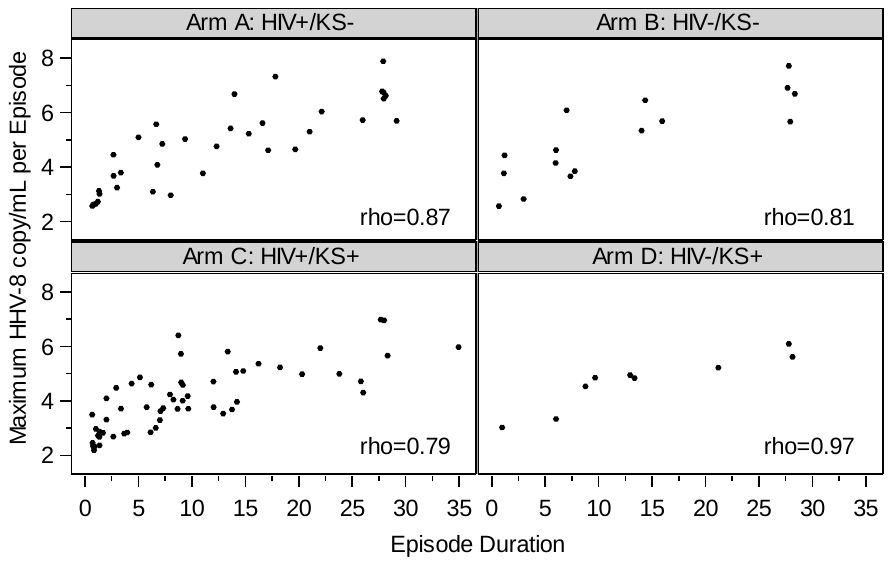

**Supplemental Figure 4**. Oral HHV-8 episode peak log_10_ copy/mL by duration, among episodes with maximum duration per participant, for each study arm.

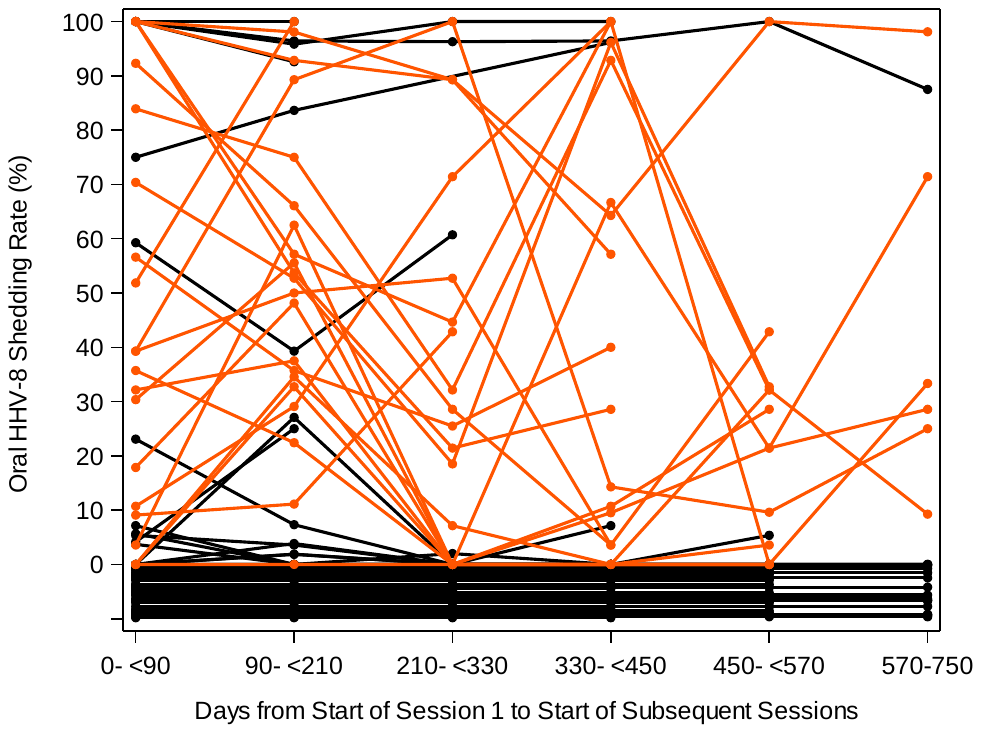

**Supplemental Figure 5.** Patterns in oral HHV-8 shedding rate over time among 92 HIV+/KS- participants contributing >1 time category. Time categories, defined relative to the start date of session 1, are shown along the x-axis. Each line represents an individual. Orange lines correspond to individuals with >30% absolute change in follow-up shedding rates compared to the initial shedding rate and black lines correspond to individuals with ≤30% change. Lines shown at or below 0% shedding rate represent individuals without HHV-8 detected in any oral swabs.

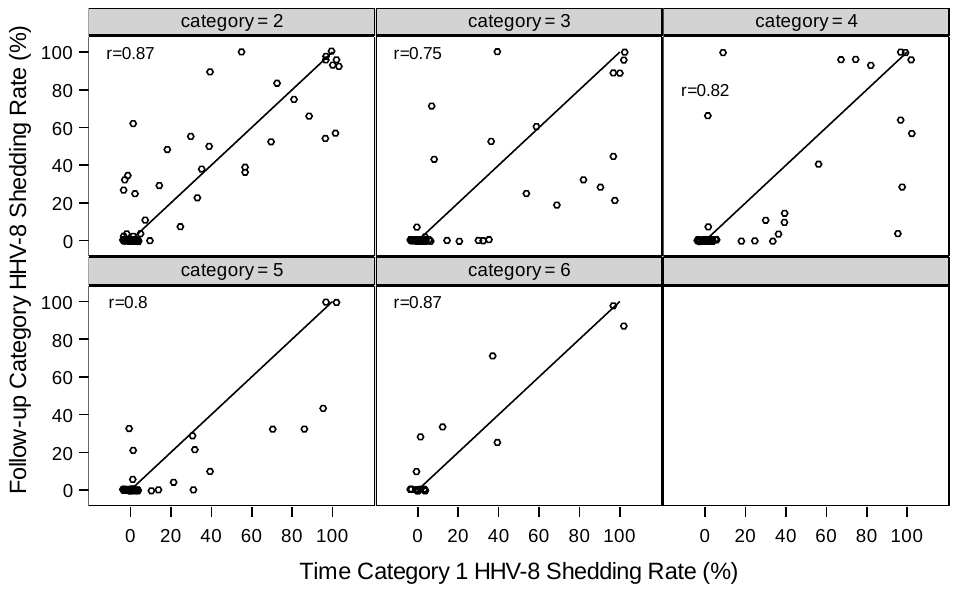

**Supplemental Figure 6.** Scatterplots for each follow-up time category oral HHV-8 shedding rate (y-axis) compared to the initial time category oral HHV-8 shedding rate (x-axis) for Arm A (HIV+ / KS-). Each panel shows data for a separate follow-up time category, with categories defined using days from the start date of the initial session to the start of subsequent sessions as follows: category 1: 0 to <90 days, category 2: 90 to <210 days, category 3: 210 to <330 days, category 4: 330 to <450 days, category 5: 450 to <570 days, category 6: 570-750 days. The line represents y=x, perfect agreement. Data have been jittered to allow viewing of overlapping data points. Spearman correlation coefficients are shown in each panel.

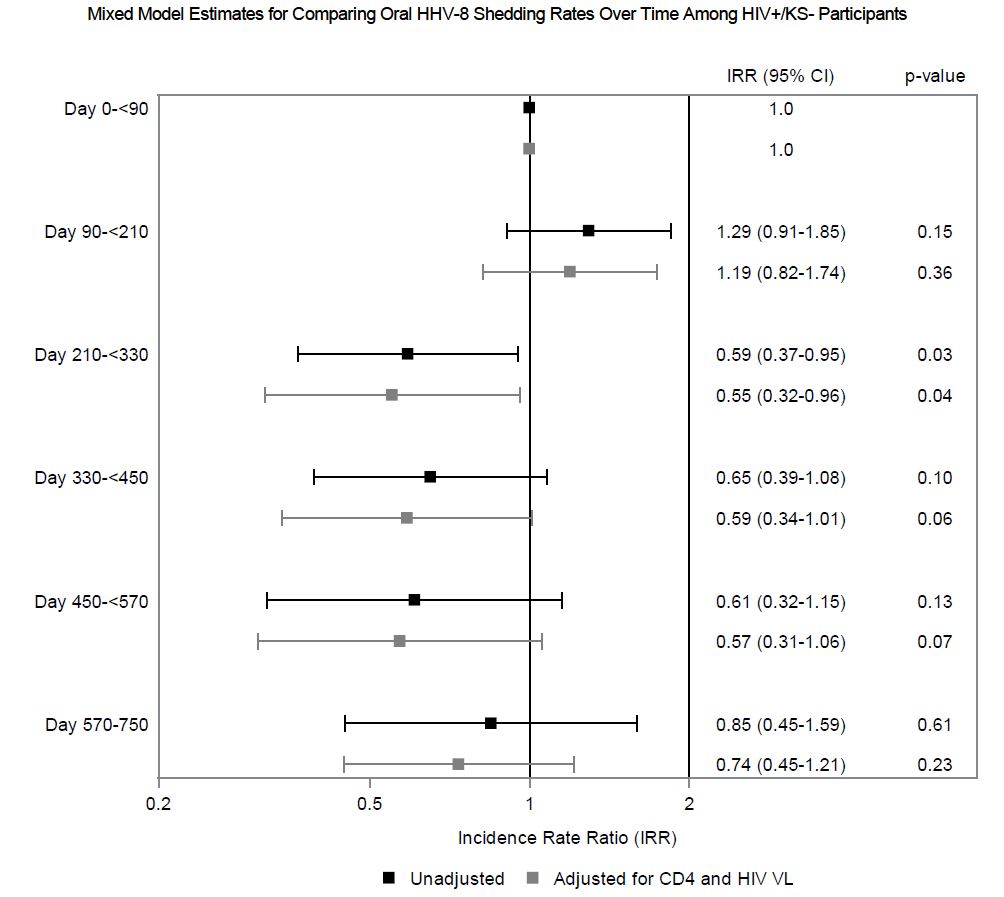

**Supplemental Figure 7.** Estimates from generalized linear mixed effects models to compare oral HHV-8 shedding rates over time among n=92 HIV+/KS- participants. Time categories are shown along the y-axis and defined by days from the start of the initial shedding session to the start of subsequent sessions. The initial time category was treated as the reference category, with each subsequent time category compared to the initial category. Model estimates are shown as incidence rate ratios (IRR) for oral HHV-8 shedding, along with 95% confidence intervals (CI). Estimates shown in black are unadjusted; estimates shown in gray are adjusted for CD4 count and HIV viral load measured at the start of each time category.

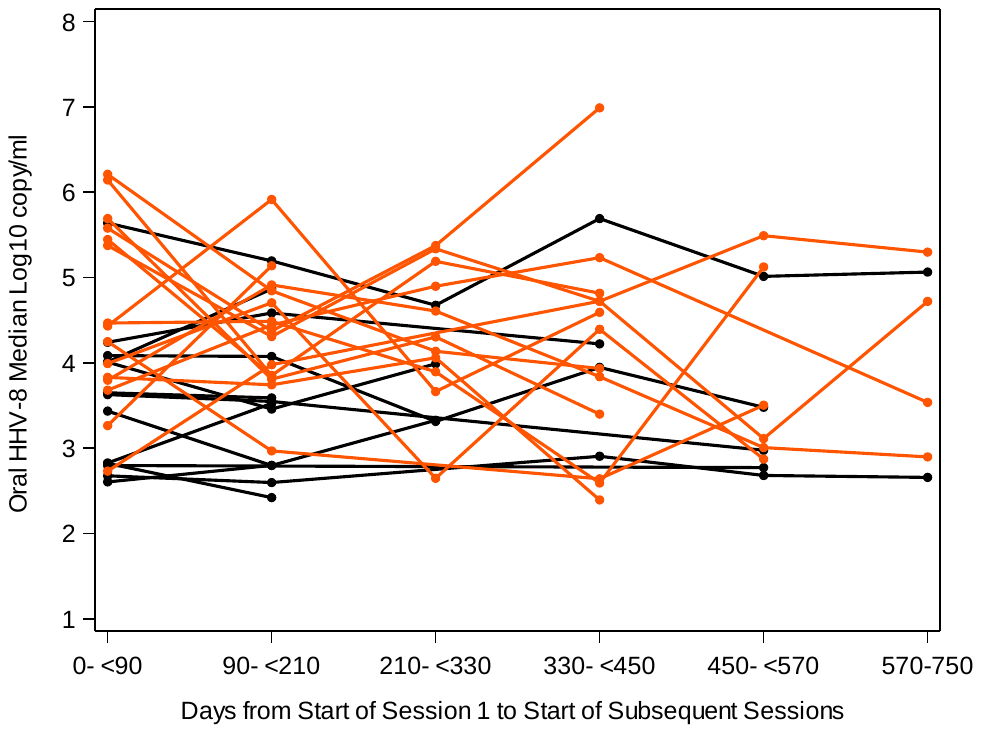

**Supplemental Figure 8.** Patterns in oral HHV-8 shedding quantity over time among 28 HIV+/KS- participants contributing >1 time category with at least one day with HHV-8 detected. Time categories, defined relative to the start date of session 1, are shown along the x-axis. Each line represents an individual. Orange lines correspond to individuals with >1 log_10_ absolute change in median HHV-8 copy number compared to the initial shedding median copy number and black lines correspond to individuals with ≤1 log_10_ change.

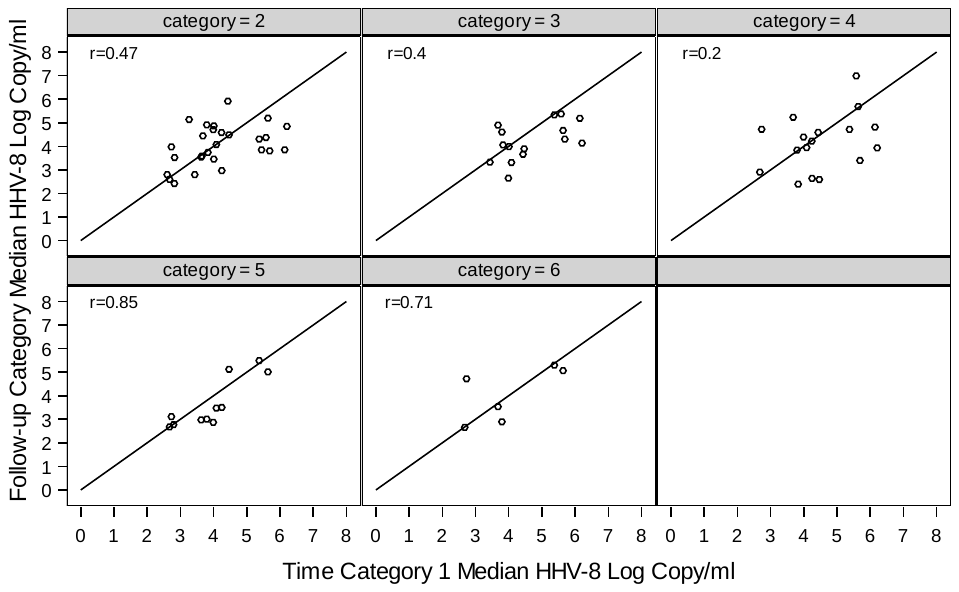

**Supplemental Figure 9.** Scatterplots for each follow-up time category oral HHV-8 median log copy per ml (y-axis) compared to the initial category oral HHV-8 median log copy per mL (x-axis) for Arm A (HIV+ / KS-). Each panel shows data for a separate follow-up category, categories defined using days from the start date of the initial session to the start of subsequent sessions as follows: category 1: 0 to <90 days, category 2: 90 to <210 days, category 3: 210 to <330 days, category 4: 330 to <450 days, category 5: 450 to <570 days, category 6: 570-750 days. The line represents y=x, perfect agreement. Spearman correlation coefficients are shown in each panel.

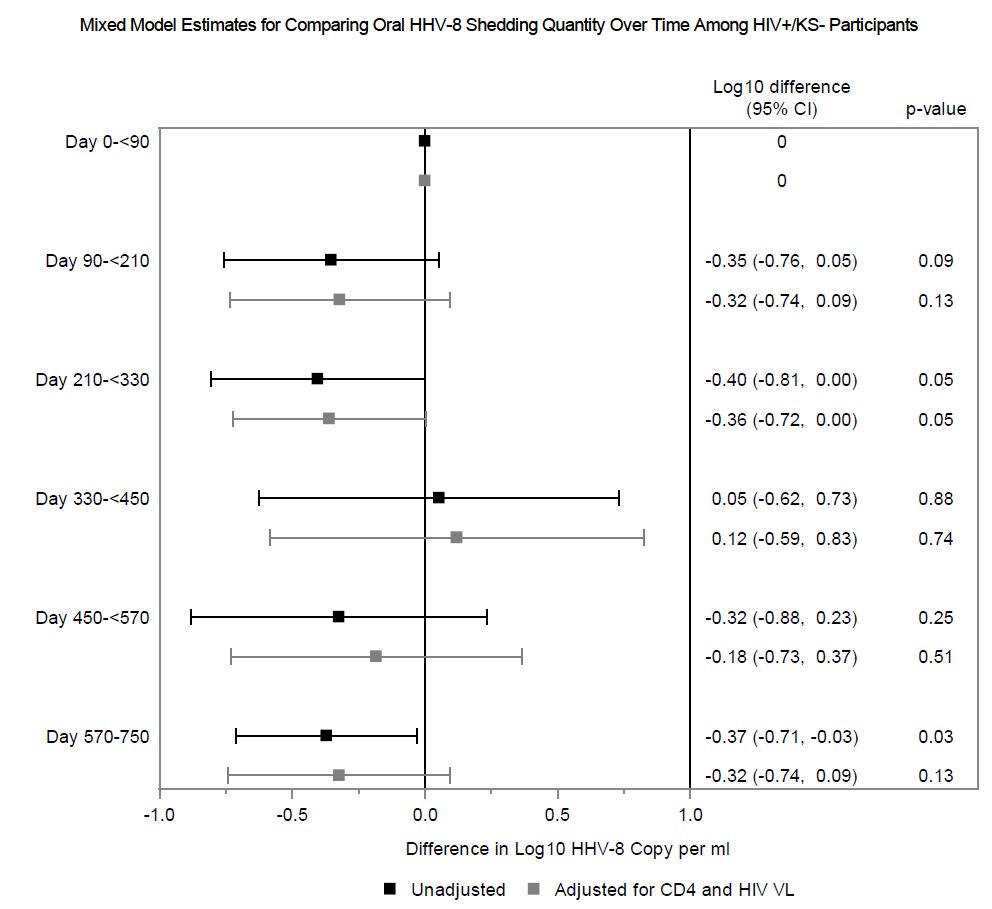

**Supplemental Figure 10.** Estimates from linear mixed effects models to compare oral HHV-8 log_10_ copy/mL over time among n=39 HIV+/KS- participants with at least one positive day and contributing data to more than one time category. Time categories are shown along the y-axis and defined by days from the start of the initial shedding session to the start of subsequent sessions. The initial time category was treated as the reference category, with each subsequent time category compared to the initial category. Model estimates are shown as mean differences in log_10_ copy/ml, along with 95% confidence intervals (CI). Estimates shown in black are unadjusted; estimates shown in gray are adjusted for CD4 count and HIV viral load measured at the start of each time category.
